# Periodontal health in a French cohort of people with Parkinson’s disease

**DOI:** 10.1101/2024.09.29.24314316

**Authors:** J Samot, E Courtin, D Guehl, N Damon-Perriere, O Branchard, M Saint-Jean, V Chuy, B Ella, MC Badet

**Affiliations:** CHU Bordeaux, Department of Oral Surgery, F-33000 Bordeaux, France; Department of Clinical Neurophysiology, Bordeaux University Hospital, Bordeaux, France. Institute of Neurodegenerative Disorders, Bordeaux; University of Bordeaux, France; Univ. Bordeaux, INRAE, Bordeaux INP, Bordeaux Sciences Agro, UMR 1366, OENO, ISVV, F-33140 Villenave d’Ornon, France; Department of Neurosurgery B, Bordeaux University Hospital, Pellegrin Hospital, 33000, Bordeaux, France; Univ. Bordeaux, INSERM, BPH, U1219, F-33000 Bordeaux, France; CHU Bordeaux, Department of Dentistry and Oral Health, F-33000 Bordeaux, France

**Keywords:** Oral Hygiene, Dental Plaque, Periodontal Index, Periodontal Diseases, Microbiota, Neurodegenerative Diseases

## Abstract

The deterioration of motor skills and fine hand movement impairment associated with Parkinson’s disease (PD) hinders the performance of daily oral hygiene procedures. This study examined the periodontal health of people with PD using clinical and microbiological periodontal parameters measured at two time points during their medical follow-up.

This prospective cohort study included participants with PD. Clinical oral health data, including oral hygiene and periodontal status, as well as microbiological samples were collected at baseline and after follow-up for at least 3 months.

Forty people with PD were recruited (35% female; median [Q1–Q3] age: 65.3 [57.9– 72.8] years) and 35 were followed up. The participants had good hygiene habits, and the proportion of participants with a “good” plaque index improved during follow-up (+21 points, *p* = 0.239). The proportion of patients with periodontal pockets > 6 mm decreased from 20% to 3% (*p* = 0.131). This improvement was associated with a reduction in the *Treponema denticola* bacterial load (*p* = 0.003).

The oral hygiene of people with PD improved during this study. Although these promising results require confirmation, they are an important step toward enhancing oral care support for PD patients.

ClinicalTrials.gov Identifier: NCT03827551, registered on January 31, 2019, (https://clinicaltrials.gov/study/NCT03827551)

## Introduction

Parkinson’s disease (PD) is a neurodegenerative disease characterized by motor and non-motor symptoms. PD affects approximately 1% of people over the age of 60 worldwide (between 6 and 7 million people in total) (de Lau and Breteler 2006; Dorsey et al. 2018). It is the second most common neurodegenerative disorder in the world after Alzheimer’s disease and the fastest growing neurological disorder worldwide (Ben-Shlomo et al. 2024; Feigin et al. 2017).

The diminished motor skills and impaired fine hand movements accompanying PD complicate daily oral hygiene procedures, and studies have shown reduced oral health and oral hygiene care in patients with PD (Einarsdottir et al. 2009; Hanaoka and Kashihara 2009; Nakayama et al. 2004; van Stiphout et al. 2018). Consequently, individuals with PD have an overall higher prevalence of oral diseases and orofacial dysfunctions, including higher prevalences of caries, gingivitis, periodontitis, and tooth loss (Bakke et al. 2011; van Stiphout et al. 2018).

Periodontal disease has a complex etiology involving microbial, inflammatory, and genetic processes and ranges clinically from simple, fully reversible gingivitis to complex forms of periodontitis. The latter, which affect the deep structures of the periodontium and destroy connective tissue and bone, result in loss of attachment, which can lead to tooth mobility and eventual tooth loss. Periodontal disease is associated with gingival recession and bleeding linked to the presence of dental plaque and calculus (Cicciu et al. 2012; Muller et al. 2011; Schwarz et al. 2006; Zarpelon et al. 2019). The progression of periodontal lesions increases with the severity of PD (Pradeep et al. 2015). At a microbiological level, the extent of periodontal damage is directly related to the total bacterial load and to the presence of bacterial species such as *Porphyromonas gingivalis*, *Aggregatibacter actinomycetemcomitans*, or *Treponema denticola* (Abusleme et al. 2021; Torrungruang et al. 2015).

Surprisingly, no studies in France have examined the oral health of people with PD, although such knowledge could enable patients to receive appropriate advice and improve their oral care to enhance their quality of life. Given the role of specific bacterial species in periodontal disease progression, analyzing the periodontal microbiota in individuals with PD can help assess potential microbial changes over time. Monitoring this variation may provide insights into periodontal health dynamics and contribute to the development of targeted preventive or therapeutic approaches.

In this context, we assessed the clinical and microbiological evolution of oral health in a group of patients with PD, with the goal of describing the periodontal health of people with PD using clinical and microbiological parameters measured at two time points during their medical follow-up.

## Material and methods

### Study design and population

This work was part of PARKIDENT, a prospective cohort study on the oral health of patients with PD (ClinicalTrials.gov identifier NCT03827551; https://clinicaltrials.gov/study/NCT03827551). The research protocol was reviewed and approved by a French regional ethics committee, the Comité de Protection des Personnes (CPP) Sud-Est III (approval number EudraCT N° 2018-A02773-52). This study adheres to the STROBE guidelines.

All participants gave informed consent. Participants were recruited between April 2019 and September 2020 at the Institute of Neurodegenerative Diseases of Bordeaux (France) according to the following inclusion criteria: have PD, older than 18 years of age, able to give informed consent, and affiliated with the national health insurance. Participants were not included if they had another condition that could affect the oral microbiome (antibiotic or antiseptic mouthwash use in the past month), if they were pregnant or wore a subtotal (fewer than six teeth with no molars or incisors) or total dental prosthesis. Including follow-up visits, data collection continued until April 2021.

### Data collection

At baseline, gender and age were recorded, along with a clinical assessment of PD. This assessment included the Movement Disorder Society–Unified Parkinson’s Disease Rating Scale (MDS-UPDRS) for motor PD symptoms (UPDRS 3), the Hoehn and Yahr scale (UPDRS 5) for PD staging, and the duration of the disease since age at diagnosis (Goetz et al. 2007; Hoehn and Yahr 1967). To address the issue of different PD drug regimens, conversion factors for antiparkinsonian drugs that have been calculated to provide a total levodopa equivalent daily dose (LEDD) were used (Tomlinson et al. 2010).

Oral health data were collected at baseline and at reassessment at least 3 months later by two dentists after a calibration phase conducted on ten patients not included in this study. The number of dental visits in the past year for baseline, or since baseline for the follow-up visit, and the toothbrushing frequency were recorded. Oral hygiene status was assessed using the Simplified Oral Hygiene Index (OHI-S) (Greene and Vermillion 1964). OHI-S is the sum of the plaque index and calculus index, which quantify the plaque and calculus on six reference teeth (four posterior and two anterior). Plaque and calculus are scored on scales from 0 to 3, and the sum of the values is divided by the number of observed surfaces. The plaque and calculus indices are interpreted as good (0–0.6), fair (0.7–1.8), and poor (1.9–3.0). The OHI-S is interpreted as good (0–1.2), fair (1.3–3.0), and poor (3.1–6.0).

The periodontal assessment included the Community Periodontal Index for Treatment Needs (CPITN) developed and validated by the World Health Organization (WHO) (Ainamo et al. 1982). A WHO basic periodontal examination probe was used. This is a lightweight metallic probe with a 0.5-mm ball tip, a black band between 3.5 and 5.5 mm, and rings 8.5 and 11.5 mm from the ball tip. The following categories were used to record periodontal status: healthy periodontium; bleeding during or after periodontal probing; presence of calculus; periodontal pocket 4–5 mm; and periodontal pocket > 6 mm.

All patients received personalized hygiene advice according to the results of the clinical examination.

### Microbiological data

Subgingival dental plaque was collected from each participant by sampling two or three of the deepest periodontal pockets among the six aforementioned reference teeth with sterile paper points, using the PERIO-ANALYSE sampling kit (Institut Clinident, Aix-en-Provence, France). Total bacterial counts were determined by the Institut Clinident using real-time quantitative PCR, including specific counts for nine bacterial species: *Aggregatibacter actinomycetemcomitans*, *Porphyromonas gingivalis*, *Tannerella forsythia*, *Treponema denticola*, *Prevotella intermedia*, *Parvimonas micra*, *Fusobacterium nucleatum*, *Campylobacter rectus*, and *Eikenella corrodens.* In addition to total bacterial quantification, the analyses by the Institut Clinident included pathogenicity thresholds initially defined by the manufacturer for the use of antibiotic therapy. Pathogenicity thresholds are expressed either as absolute numerical values or as natural logarithms (ln).

### Statistical analyses

Analyses were performed with R (v 4.2.1); the level of statistical significance was set at *p* < 0.05 (R Core Team 2020). Quantitative variables were described as means, standard deviation, medians, first and third quartiles, and range. Qualitative variables were described as frequencies and percentages. Paired comparisons between the baseline and follow-up examinations were made using the Wilcoxon signed-rank test and McNemar’s test.

## Results

### Demographic data

At the time of study design, no data were available to calculate the sample size. Therefore, the sample size was determined based on our recruitment capacity, with a maximum inclusion period of 12 months. This study included 40 patients with PD (65% male; median age 65.3 [range 39.4–81.1] years) (Table 1). At baseline, the median disease duration was 14.5 years. The median UPDRS 3 score was 14.5 and the median UPDRS5 was 2, indicating a good degree of autonomy.

**Table 1:** Participant characteristics at baseline.

| Characteristics at baseline | TOTAL (N=40) |
| --- | --- |
| **Female** | 14 (35) |
| **Age** (years) | 65.3 [57.9-72.8] |
| **UPDRS3 Score**, 2 m.d. | 14.5 [10.0-23.5] |
| **UPDRS5 Score**, 2 m.d. | 2.0 [2.0-3.0] |
| **Duration of the disease** (years), 2 m.d. | 14.5 [9.0-20.0] |
| Values in N (%) for the qualitative variable and in median [1st quartile – 3rd quartile] for quantitative variables. |  |
| m.d.: missing data; UPDRS: Unified Parkinson’s Disease Rating Scale. |  |

The follow-up visit was disrupted by the COVID-19 crisis. The median time to follow-up was 7 [range 3–21] months. Thirty-five patients completed a follow-up visit, and five withdrew from the study for health reasons (*e.g.*, broken femur, fatigue, deterioration in general condition).

### Levodopa equivalent daily dose (LEDD)

At baseline, the mean ± SD LEDD was 784.2 ± 638.7 mg/d, and a non-significant decrease was observed at follow-up (778 ± 597.4 mg/d, *p* = 0.96). One patient had missing data at both baseline and follow-up.

### Oral care habits

In total, 37.5% of the participants reported no dental examinations in the previous year (Table 2). Among those with regular dental appointments, 16% reported three or more dental visits in the previous year. All reevaluated patients reported at least one dental visit since baseline.

**Table 2:** Participants’ oral care habits.

| Characteristics | Baseline<br>(N=40) | Follow-up visit<br>(N=35) | p* |
| --- | --- | --- | --- |
| **Dental visit in the past year** | 25 (62) |  |  |
| **Number of visits to the dentist in the past year** (for those with $\geq 1$ visit) | 1.0 [1.0-2.0] | | n.a. |
| 1 | 16 (40) |  |  |
| 2 | 5 (13) |  |  |
| $\geq 3$ | 4 (10) | | |
| **Number of visits to the dentist since baseline, 3 m.d.** |  |  | n.a. |
| None |  | 10 (29) |  |
| 1 |  | 19 (54) |  |
| 2 |  | 3 (9) |  |
| **Toothbrushing frequency, 1 m.d.** |  |  | 0.134 |
| <2 times/day | 12 (30) | 7 (20) |  |
| $\geq 2$ times/day | 27 (68) | 27 (77) | |
| **Plaque index, 1 m.d. at follow-up** |  |  | 0.239 |
| Good | 20 (50) | 25 (71) |  |
| Fair | 20 (50) | 9 (26) |  |
| **Calculus index, 1 m.d. at follow-up** |  |  | 0.248 |
| Good | 35 (88) | 31 (89) |  |
| Fair | 5 (13) | 3 (9) |  |
| **OHI-S, 1 m.d. at follow-up** |  |  | 0.121 |
| Good | 27 (68) | 30 (86) |  |
| Fair | 13 (33) | 4 (11) |  |
| **CPITN status** |  |  | 0.131 |
| Healthy periodontium | 10 (25) | 8 (23) |  |
| Bleeding during or after periodontal probing | 3 (8) | 6 (17) |  |
| Presence of calculus | 4 (10) | 10 (29) |  |
| Periodontal pocket 4-5 mm | 15 (38) | 10 (29) |  |
| Periodontal pocket >6 mm | 8 (20) | 1 (3) |  |
| **CPITN status for posterior teeth** |  |  | 0.091 |
| Healthy periodontium | 11 (28) | 12 (34) |  |
| Bleeding during or after periodontal probing | 7 (18) | 10 (29) |  |
| Presence of calculus | 2 (5) | 3 (9) |  |
| Periodontal pocket 4-5 mm | 13 (33) | 9 (26) |  |
| Periodontal pocket >6 mm | 7 (18) | 1 (3) |  |

Values in N (%) for qualitative variables and in median [1st quartile – 3rd quartile] for the quantitative variable.
m.d.: missing data; CPITN: Community Periodontal Index for Treatment Needs; n.a.: not applicable; OHI-S: Simplified Oral Hygiene Index.
\* McNemar's Chi-squared test with continuity correction.

Most participants reported brushing their teeth twice a day at baseline, and a non-significant increase in the frequency was observed at the follow-up visit (68% *vs*. 77%, *p* = 0.134). There was also an improvement in the plaque index between baseline and the follow-up visit, whereas the calculus index remained stable. Therefore, the OHI-S improved (68% *vs*. 86% with good oral hygiene, *p* = 0.121).

Regarding periodontal status, there was a decrease in the frequency of patients with healthy periodontium, but also a decrease in the frequency of patients with periodontal pockets, with 20% at baseline of patients having periodontal pockets > 6 mm versus 3% follow-up (overall, *p* = 0.131 for CPITN status at baseline *vs*. follow-up). The positive trend in pocket depth was marked for posterior teeth.

### Microbiological data

Thirty-three patients had data on the quantity of the nine assessed bacteria at both baseline and follow-up (Table 3). The median bacterial load decreased significantly between baseline and follow-up only for *T*. *denticola* (*p* = 0.003), dropping from a pathogenic threshold to a physiological threshold. For *A*. *actinomycetemcomitans*, the bacterial load was zero at both baseline and follow-up, indicating quantities below the PCR detection threshold. For the other bacteria and the total bacterial load, the quantities remained similar between baseline and follow-up.

**Table 3:** Description of periodontal bacteria in patients at baseline and at the follow-up visit (ln-transformed)

| Periodontal bacteria | Pathogenicity threshold | Baseline<br>(N=33) | Follow-up visit<br>(N=33) | p |
| --- | --- | --- | --- | --- |
| Total Flora | Healthy periodontium < 16.1<br>Gingivitis < 18.4<br>Periodontitis > 18.4 | 21.6 [20.7-22.6] | 21.4 [20.8-22.5] | 0.406 |
| *Aggregatibacter actinomycetemcomitans* | 6.9 | 0.0 [0.0-0.0] | 0.0 [0.0-0.0] | 0.789 |
| *Porphyromonas gingivalis* | 11.5 | 10.9 [0.0-14.5] | 10.4 [0.0-12.0] | 0.338 |
| *Tannerella forsythia* | 11.5 | 12.8 [11.4-14.0] | 11.8 [10.0-14.5] | 0.660 |
| *Treponema denticola* | 11.5 | 13.4 [10.9-15.8] | 11.1 [8.6-13.3] | **0.003** |
| *Prevotella intermedia* | 11.5 | 16.6 [13.9-17.5] | 16.5 [13.9-17.4] | 0.427 |
| *Parvimonas micra* | 13.8 | 14.1 [12.5-15.2] | 13.5 [11.6-15.4] | 0.235 |
| *Fusobacterium nucleatum* | 16.1 | 15.8 [15.3-17.0] | 15.9 [13.8-17.0] | 0.321 |
| *Campylobacter rectus* | 13.8 | 14.6 [13.5-15.5] | 13.2 [11.4-15.0] | 0.065 |
| *Eikenella corrodens* | 16.1 | 13.1 [11.5-14.3] | 11.4 [8.2-14.0] | 0.173 |
| Values in median [1st quartile – 3rd quartile]. |  |  |  |  |

At baseline and follow-up, more than 70% of the patients had total bacterial counts consistent with the presence of periodontitis (Table 4), whereas the proportion of patients with a bacterial load compatible with gingivitis decreased from 27% at baseline to 18% at follow-up.

**Table 4:** Description of periodontal bacteria at baseline and at the follow-up visit in relation to pathogenicity thresholds.

| Periodontal bacteria | Baseline<br>(N=33) | Follow-up visit<br>(N=33) | p |
| --- | --- | --- | --- |
| **Total flora** |  |  | 0.456 |
| Healthy periodontium | 0 | 1 (3) |  |
| Gingivitis | 9 (27) | 6 (18) |  |
| Periodontitis | 24 (73) | 26 (79) |  |
| *A. actinomycetemcomitans* $\geq 1e+0.3$ | 3 (9) | 3 (9) | 1 |
| *Porphyromonas gingivalis* $\geq 1e+0.5$ | 15 (46) | 12 (36) | 0.546 |
| *Tannerella forsythia* $\geq 1e+0.5$ | 24 (73) | 18 (55) | 0.211 |
| *Treponema denticola* $\geq 1e+0.5$ | 23 (70) | 15 (46) | 0.080 |
| *Prevotella intermedia* $\geq 1e+0.5$ | 31 (94) | 29 (88) | 0.480 |
| *Parvimonas micra* $\geq 1e+0.6$ | 18 (55) | 15 (46) | 0.803 |
| *Fusobacterium nucleatum* $\geq 1e+0.7$ | 14 (42) | 16 (49) | 0.814 |
| *Campylobacter rectus* $\geq 1e+0.6$ | 22 (67) | 15 (46) | 0.121 |
| *Eikenella corrodens* $\geq 1e+0.7$ | 2 (6) | 2 (6) | 1 |
| Values in N (%). |  |  |  |

## Discussion

In this study, the participants tended to maintain good oral hygiene habits, as reflected in both toothbrushing frequency and plaque levels at baseline and follow-up. Over time, a marked reduction in pocket probing depth (PPD) was observed, particularly for depths greater than 6 mm. Additionally, there was a significant decrease in *T*. *denticola* counts, along with reductions in the total bacterial load and the load of six other bacterial species. However, despite these improvements, more than 7 in 10 patients continued to have bacterial counts consistent with periodontitis throughout the study.

The toothbrushing frequency observed in this study was similar to that reported in other studies for populations with PD (Einarsdottir et al. 2009; Lyra et al. 2020; Verhoeff et al. 2022). According to Einarsdottir et al., who investigated individuals with PD in Iceland, these levels were even higher than those observed in a control population, although the difference was not significant (Einarsdottir et al. 2009). Notably, these results were obtained despite the absence of a specific dental care policy for people with PD in Iceland at the time.

Although our study participants appeared to follow toothbrushing recommendations, many still had plaque accumulation, suggesting ineffective brushing techniques. At follow-up, after all participants reported having consulted a dentist, plaque levels improved, possibly reflecting a positive impact of these consultations or even participation in the study itself. These findings align with those of Baram et al., who reported a significant reduction in the plaque index 2 months after an intervention in two groups of approximately 15 patients (Baram et al. 2020).

The assessment of calculus presence is more complex due to the use of two different indices in this study: the calculus index integrated into the OHI-S, and the CPITN. Based on the OHI-S alone, the presence of calculus was similar between baseline and follow-up, whereas participants’ oral hygiene appeared to improve as determined by plaque levels. When assessed using the CPITN, calculus appeared more prevalent among patients who attended the follow-up visit. Nevertheless, the CPITN analysis also revealed a marked reduction in periodontal pockets exceeding 4 mm in depth, particularly in posterior teeth. This trend suggests potential regression in the severity of periodontal damage, despite the persistence of calculus. Although plaque levels decreased over time, the continued presence of substantial calculus can be explained by the lack of scaling between baseline and follow-up. Since calculus forms from mineralized plaque that has not been entirely removed, its persistence is expected. Scaling is a professional procedure requiring a dentist, whereas plaque removal can be performed at home.

Consistent with these findings, the need for periodontal care was high at enrollment due to the presence of deep periodontal pockets, but improved over time. Overall, these data suggest that despite multiple risk factors for poor oral health, patients with PD actively engage in maintaining good oral hygiene. To our knowledge, only one randomized controlled trial has evaluated the effect of an intervention on oral health parameters in PD patients (Baram et al. 2020; Baram et al. 2021).

Among studies assessing PPD in individuals with PD, few have used the CPITN, and those that have tend to report only a global score, which limits insight by failing to differentiate between individual CPITN components (Barbe et al. 2017; Garcia-de-la-Fuente et al. 2023). However, other studies have highlighted increased pocket depth in PD patients. Cicciu et al. reported that 75.8% of cases presented with pocket depths greater than 5 mm, whereas Hanaoka and Kashihara found that 99% of PD patients had pockets exceeding 4 mm, compared to 44% in their control group (Cicciu et al. 2012; Hanaoka and Kashihara 2009). In our study, deep periodontal pockets were prevalent in the posterior dental region. When combined with tooth mobility, this can cause pain and significantly impair masticatory function. Moreover, increased pocket depth in molars ultimately worsens their long-term prognosis for retention in the dental arch (Pereira et al. 2012).

Interestingly, despite persistent calculus, PPD improved over time. Previous studies have shown that gingival health can improve in the presence of significant calculus deposits, provided that plaque levels are reduced (Gaare et al. 1990). This underscores the crucial role of microbial control in the progression of periodontal disease.

The microbiological data further emphasize a precarious periodontal state, with five of the nine bacteria studied having median values above defined pathogenicity thresholds and a bacterial load incompatible with periodontal health. Limited data exist on the composition and abundance of periodontal disease-associated bacterial flora in individuals with PD. In a cohort of about 20 patients, Yay et al. observed that the subgingival microbiome associated with periodontitis was altered in PD patients. In deep periodontal pockets, *P*. *micra*, *C*. *rectus*, and *T*. *denticola*, among others, were detected more frequently in individuals with PD compared to those with periodontitis in the general population (Yay et al. 2023). Despite follow-up, many patients retained high bacterial loads, with only one exhibiting a total bacterial load compatible with good periodontal health. These findings align with those of He et al., who demonstrated that PPD correlates poorly with the total bacterial load, but is more strongly associated with the presence of specific pathogens such as *P*. *gingivalis*, *T*. *forsythia*, and *T*. *denticola* (He et al. 2020). In our study, the *T. denticola* load significantly decreased over time. This reduction may reflect an improvement in plaque levels, but could also result from factors that have not yet been fully identified. Some authors have suggested that catecholamines, including dopaminergic agents, influence bacterial load and ultimately dental health (Jentsch et al. 2013; Roberts et al. 2002). However, it is unlikely that the observed changes in our study were related to these agents, as the mean LEDD remained relatively stable throughout the study. Addressing the bacterial origins of inflammation in periodontitis is crucial, as numerous studies suggest that periodontal lesions contribute to systemic inflammation, potentially exacerbating neuroinflammation (Adams et al. 2019; Hajishengallis 2015; Li et al. 2022). This study is also of interest from a regional perspective, as geographical variation in the presence of bacterial species associated with severe periodontal disease has been reported (Lafaurie et al. 2022). Further studies incorporating more extensive microbiological data are therefore needed.

This study enrolled a small sample recruited from one hospital, which limits the generalizability of the findings to other populations. However, its longitudinal design is a strength, enabling the assessment of clinical and microbiological periodontal parameters over time—including their maintenance, deterioration, or improvement—despite disruptions caused by the COVID-19 crisis. All participants also underwent clinical evaluations for both PD and oral health, and the use of validated indices alongside biological parameters likely minimized information bias.

Our findings highlight a positive trend in oral hygiene among individuals with PD included in this study. Although these promising results require validation, they are an important step toward strengthening oral care support for PD patients. However, both our study and previous ones focused primarily on patients with relatively mild PD. Future research should extend these findings by including individuals with more advanced disease.

Finally, this study, along with recent research on the oral health of PD patients, contributes to the development of tailored recommendations for their multidisciplinary management. By expanding the literature on oral health in PD, our findings reinforce existing recommendations, such as those summarized by Martimbianco et al., which outline key strategies to improve the quality of life of individuals with Parkinson’s disease (Martimbianco et al. 2021).

## Data Availability

All data produced in the present study are available upon reasonable request to the authors

## Acknowledgments

We would like to express our sincere gratitude to Drs. Magnim Bassou and Cécilia Dapremont. As dental students at the time of data collection, they provided invaluable assistance in patient follow-up.

Martine Saint-Marc, a biomedical research technician, is gratefully acknowledged for technical assistance in the laboratory.

Our last words are dedicated to the memory of the recently deceased Dr. Marie-Cécile Badet, who was one of the investigators of the team and contributed to the genesis of this project.

This study was supported by France Parkinson.

The authors declare no potential conflicts of interest.

